# Rest-Activity Rhythm Variability Across Clinical Episodes of Bipolar Disorder: Standalone Biomarker or Statistical Artifact?

**DOI:** 10.64898/2026.07.15.26358139

**Authors:** Carmen-Anna Konicarova, Jakub Schneider, Marian Kolenic, Filip Spaniel, Martin Alda, Eduard Bakstein

## Abstract

**Background:** Actigraphy-derived rest–activity rhythm (RAR) features are widely used to characterize clinical states in bipolar disorder (BD). Both mean levels and temporal variability of these features have been associated with mood episodes; however, variability measures are often statistically coupled with the mean, particularly in skewed distributions. This raises a question as to whether variability reflects a separate characteristic of the data or whether the observed association arises from statistical properties of the data.

**Objective:** In this study, we aim to determine whether temporal variability of actigraphy-derived RAR features provides standalone information on mood episodes in BD beyond mean activity levels after accounting for mean–variance dependence.

**Methods:** We analyzed actigraphy data from a subset of 72 participants with BD drawn from a larger longitudinal study, extracting 22 daily RAR features aggregated weekly as sample mean (MEAN) and within-week temporal variability computed as sample standard deviation (VAR). Variance-stabilizing transformations (Box–Cox or Yeo–Johnson) were applied to the entire study cohort to reduce mean–variance dependence. Associations with mood episodes and remission (mania: n=34; depression: n=58 annotated participants) were evaluated using generalized linear mixed-effects models with a logistic link function, including univariate (MEAN or VAR) and multivariate (MEAN+VAR) specifications, assessed by likelihood-based metrics and the area under the receiver operating characteristic curve (AUC).

**Results:** Transformations reduced mean–absolute correlations from 0.43 to below 0.06. Temporal variability remained significantly associated with clinical state for 11/22 RAR features in mania and 16/22 features in depression, with all significant associations remaining after false discovery rate correction (p<0.05). Joint models showed modest incremental gains (AUC 3%–4% overall; up to 12% in mania, 7% in depression), with absolute performance remaining limited (AUC 0.50–0.66). In both mania and depression, nearly all significant variability-based regressors contributed incremental information beyond mean-based models. Only sleep duration and activity changes around wake time (±1 hour), did not improve discrimination between mania and remission.

**Conclusions:** Temporal variability in RAR features can be considered a standalone state marker of mood episodes not captured by mean activity. We found it to be more consistently associated with depression than mania. Its incremental discriminative contribution is modest, suggesting greater utility within multivariate or multimodal frameworks.

**Key strengths:**

- **Large-scale longitudinal design:** The analysis leverages 76,672 days of actigraphy data from 326 participants with BD, enabling robust estimation of within-participant dynamics and increasing statistical power. Analysis sample sizes were smaller for specific contrasts (mania–remission: 34 participants; depression–remission: 58 participants), reflecting the longitudinal nature of bipolar disorder (BD), sparseness of episodes, and the challenges of acquiring dense actigraphy data in clinical populations.
- **Direct enumeration of mean–variance coupling:** The study explicitly quantifies and mitigates mean–variance dependence using feature-wise variance-stabilizing transformations, addressing a critical but often neglected methodological issue, which has broader biomedical applications.
- **Temporal variability is a standalone state marker in BD:** After transformation, temporal variability was significantly associated with clinical state in the majority of features (mania: 11/22; depression: 16/22), indicating that these effects were not solely attributable to statistical coupling with the mean, and that variability of RAR is an inherent biomarker.

## Introduction

The clinical course of BD is characterized by mood episodes associated with changes in energy and activity, as well as disruptions of circadian rhythms [1, 2]. Actigraphy offers an objective and non-invasive method for quantifying rest-activity-rhythms (RARs) in real-life conditions [3]. Therefore, it allows continuous and long-term monitoring, making it particularly suitable for studying clinical conditions in BD patients [4, 5], and can help assess activity patterns in different phases of the illness.

### Rest-Activity Findings in Patients with Bipolar Disorder

Actigraphy-based assessments of RARs have become an important objective tool for characterizing mood states in BD. A consistent finding across studies is that mean activity levels reliably differentiate affective polarity: manic episodes are associated with elevated activity energy expenditure relative to euthymic states [6, 7, 8], whereas depressive episodes are characterized by reduced overall activity [6, 9, 10, 8]. Euthymic states typically fall intermediate between these two extremes, although residual abnormalities in activity regulation persist even during symptom remission [11].

Despite their discriminative value, mean-based measures show limited utility in predicting imminent mood transitions. Longitudinal analyses indicate that changes in average activity levels are not consistently directionally aligned prior to episode onset and therefore fail to serve as reliable early warning indicators [12, 13]. This limitation has shifted attention toward variability– and complexity-based features of RARs.

Variability metrics capture dynamic instability that is not reflected in mean activity alone. Across studies, manic states have been associated with reduced short-term variability in activity counts, alongside increased signal complexity and rhythm fragmentation [12, 14, 15]. This apparent dissociation suggests that manic behavior may be characterized by rapid alternations within an overall dysregulated but structured circadian pattern. In contrast, depressive episodes exhibit a different profile, characterized by reduced day-to-day rhythm stability together with more rigid and stereotyped within-day activity patterns [6, 15]. Together, these findings suggest that depression reflects a constrained expression of behavior within a weakened or phase-shifted circadian framework.

Empirical evidence further supports the superiority of variability-based measures over mean activity in detecting state instability. Reduced standard deviation combined with increased temporal complexity (e.g., sample entropy) has been observed in manic states compared to remission [12], and entropy has been shown to discriminate between clinical states [16]. Similarly, increased minute-to-minute variability has been reported in clinically unstable euthymic individuals, suggesting persistent trait-like dysregulation even in the absence of acute episodes [17]. Importantly, early warning signal approaches based on variability dynamics have demonstrated predictive capability for mood transitions up to four weeks in advance, outperforming mean-based indicators [13].

Longitudinal digital phenotyping studies provide additional granularity regarding temporal variability patterns preceding episode onset. Wearable-derived sleep, circadian, and activity features have demonstrated predictive value for mood episode onset in BD, with machine learning approaches achieving meaningful discrimination between mood states [18, 19, 20, 21, 22, 23]. Importantly, variability-based and complexity-based features have shown particular promise for predicting mood transitions with clinically relevant lead times, in some cases preceding episode onset by several weeks [13, 12, 24, 25]. At the feature level, increased day-to-day variability in step count preceded depressive symptom onset by a median of 7 days in a cohort of 127 outpatients with BD, suggesting that heightened instability may serve as an early marker of depressive relapse [26]. In contrast, multimodal behavioral data from 133 individuals with BD identified reduced variability in mood and activity, alongside increased variability in sleep onset latency, as key predictors of depressive episodes [27]. These seemingly divergent findings likely reflect differences in operational definitions of variability and the temporal scale of measurement. In relation to manic transitions, emerging evidence suggests that alterations in sleep–wake dynamics may precede changes in motor activity. Specifically, within-night variability in sleep architecture has been identified as an early indicator of manic onset, with activity-based changes emerging subsequently while still contributing meaningful predictive information [28].

### Methodological Considerations and Current Gaps

Actigraphic studies in BD show that measures of variability tend to align more closely with mood states than mean-based measures [25, 29, 12, 26, 28].

However, an important concern is that variability measures are usually statistically coupled with mean values, particularly in skewed distributions. For instance, the observed increase in variability during manic phases, particularly within skewed distributions of mean activity between 00:00 and 06:00 local time, may be confounded by concurrent shifts in central tendency and distributional shape, rather than representing a true increase in intrinsic variability of nocturnal activity patterns. Despite the frequent use of both parameters in BD research, the relationship between mean and variability remains in most cases insufficiently clarified.

Partial independence of variability and mean measures is suggested by several earlier findings: day-to-day variability in activity detected depressive symptoms 7 days earlier than changes in mean sleep duration [26], adding intra-day variability to clinical predictors improved classification of early versus late recurrence from area under the receiver operating characteristic curve (AUC) 0.64 to 0.82 [25], and stepwise regression retained both mean measures (sleep duration, sleep latency) and variability measures (fragmentation index variability) in optimal models achieving 89% classification accuracy for euthymic versus depressive states [30].

### Objectives of the Present Study

We analyzed data from a large longitudinal actigraphy study to examine the relative role of variability in RAR actigraphic features as a potential biomarker of mood states and relapse status in BD, and to what extent it provides additional information over the mean-based features. In other words, do variability measures represent independent physiological information or reflect statistical coupling with the mean (i.e., mean–variance collinearity)?

The study addresses the following research questions:

- **Q1:** Is the association of variability of RAR measures with mood states in BD explained by correlation with the mean measures?
- **Q2:** Which RAR variability-based features distinguish mood states in BD when mean–variance coupling is reduced?
- **Q3:** Does RAR variability in actigraphy features add information to the mean measures to better identify relapse status or mood state transitions in BD?

To disentangle these effects, we applied a deskewing transformation to reduce mean–variance association and to isolate the component of variability that is minimally dependent on the mean. This approach allows us to assess whether variability in RAR features contributes unique in-sample information regarding clinical outcomes in BD. In this study, a ‘standalone regressor’ or ‘standalone marker’ is defined as an input variable that retains a statistically significant association with the outcome after controlling for mean–variance coupling.

## Data and Methods

### Participants and Data

This study utilized data collected from a previously conducted observational, non-interventional study AKTIBIPO 400 [31]. The study cohort comprised 326 participants with BD, aged 18–65 years, who were euthymic at baseline. The diagnosis of BD was established according to ICD-10 criteria by the treating psychiatrist before study inclusion. Participants were followed for mean 235 ± 214 days with MindG continuous wrist-worn actigraphy by Mindpax Ltd., they self-reported manic and depressive symptoms weekly using online assessments by the digital questionnaire Aktibipo Self-rating Ecological Momentary Assessment (ASERT) by Mindpax Ltd. [31], a brief 10-item self-assessment scale for manic and depressive symptom severity, validated for digital monitoring [32, 33]. A subset of 115 participants additionally underwent monthly clinician-rated Montgomery-Åsberg Depression Rating Scale (MADRS) and Young Mania Rating Scale (YMRS) evaluations through telephone interviews.

The study was approved by the NIMH institutional ethical committee (Czech Republic; case number: 101/17). Upon enrollment, all participants signed informed consent, and the study was conducted in compliance with the Declaration of Helsinki.

### Clinical Assessment and Episode Definition

In this work, we sought to identify differences in actigraphy-derived features between clinical states. The clinical states were annotated using a rule-based algorithm using details from three primary sources: monthly clinical assessments (MADRS, YMRS), weekly self-reported questionnaire (ASERT) including depressive and manic subscores, and health records—containing data on hospital admissions and sick leave associated with affective symptoms.

Episode classification relied primarily on scores from MADRS and YMRS. A (hypo)manic episode was assigned if YMRS ≥ 13 in a 14-day period or if mania-related entries appeared in health records. A depressive episode was identified within a 14-day interval if MADRS ≥ 15 or if depression-related events were recorded in health records. For both episode types, classification was extended to adjacent weeks when the corresponding ASERT subscore or its weekly change surpassed a predefined threshold. Remission was defined by MADRS and YMRS values *<* 9, with ASERT depressive subscore *<* 5 and manic subscore *<* 3. Short periods of missing data (≤ 11 days) were filled in by maintaining the same state if it was consistently observed before and after the gap.

The daily annotated dataset (115 participants) was stratified into two binary subsets: mania–remission and depression–remission. Participants were included in a subset if they had at least one week classified as the respective episode state and at least one week classified as remission, according to the criteria outlined above.

### Actigraphy and Daily Features

Continuous wrist actigraphy was collected and aggregated into 30-second epochs. From these data, 22 daily features were derived, comprising 8 daytime and 14 sleep-related features. Table 2 provides a detailed overview of all computed actigraphic features. Due to the study design, feature extraction was limited to the same-day (night) data. Therefore, actigraphy metrics requiring data from multiple adjacent days, such as cosinor analysis, were excluded.

### Actigraphic Weekly Aggregates

Weekly temporal aggregation of the actigraphy features was performed using two summary statistics: mean-based approach (MEAN) computed as the sample mean and time-variability-based approach (VAR) computed as standard deviation in weekly window. Aggregations were computed separately for the mania–remission and depression–remission subsets. Only participants who experienced at least one symptomatic episode and at least one remission during the observation period were included in the corresponding subset. To ensure the reliability of the aggregated clinical labels, aggregation windows were retained only if they contained at least four days with consistent clinical annotation.

Daily actigraphic data consisted of selection of features (*n* = 22) measured for each day of each participant. Then, for each participant (*i*), each feature (*j*) was aggregated within non-overlapping 7-day windows (*W* = 7) in order to reduce the influence of the circaseptan rhythm, defined as:

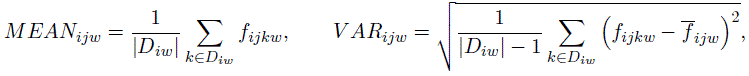

where *D_iw_* denotes the set of valid days within week *w* for participant *i*, and |*D_iw_*| is its cardinality (i.e. the number of valid values), *f_ijkw_* denotes the value of actigraphic feature (*j*) for each participant (*i*) on day (*k*) of week *w*. The quantity *MEAN_ijw_* represents the weekly sample mean of valid days of given feature, while *V AR_ijw_* represents the within-week temporal variability measured as the sample standard deviation of valid days.

### Power-Transformation–Based Control of Mean–Variability Dependence

To address whether the variability of an actigraphy feature constitutes a standalone regressor, or whether its apparent effect is primarily driven by collinearity with the mean, we constructed two variants of the dataset (each comprising 326 participants and 76,672 days to maximize the robustness of the estimated transformation): a) the original dataset, and b) a power-transformed dataset in which both the Box-Cox power-transformation (B-C) [34] and Yeo-Johnson power-transformation (Y-J) [35] were applied to each of the 22 daily actigraphy features. For each feature, we selected the transformation (B-C or Y-J) that resulted in lower absolute correlation between its MEAN-VAR weekly aggregates in each subset (mania or depression, see section Actigraphic Weekly Aggregates). This criterion was chosen as a pragmatic proxy for minimizing mean–variance coupling, although it does not guarantee full statistical independence. Whereas Y-J is, by definition, applicable to both positively and negatively skewed data and can also process negative values [35], B-C tends to fail on data with left-skewed distributions, and needs the data to be positive [34].

All subsequent preprocessing and aggregation steps described above for the original dataset were also applied identically to the power-transformed variant of the dataset.

### Statistical Modeling

To address research questions: ‘Is RAR variability in actigraphy features only a technical artifact?’ (Q1) and ‘Does RAR variability in actigraphy features distinguish clinically relevant mood states in BD when mean–variance coupling is reduced?’ (Q2), we employed univariate generalized linear mixed-effects models (GLMMs). To address the concerns regarding the incremental explanatory information of variability beyond the mean (Q3), we fitted multivariate models including both MEAN and VAR simultaneously using the transformed data.

The analytical sample comprised 72 participants who met the inclusion criteria for at least one analysis. Specifically, the mania–remission models were fitted using data from 34 participants, whereas the depression–remission models were fitted using data from 58 participants.

All models (Q1-Q3) were specified as GLMMs with a binomial distribution and logit link function. Parameters were estimated via maximum likelihood using the Laplace approximation, and observation-level weights were incorporated to account for class imbalance in the clinical labels. Each observation corresponded to a weekly aggregation of a specific actigraphic feature (either MEAN or VAR). Time was defined as the date corresponding to the final day of the aggregation window. The binary clinical outcome (episode vs. remission) was used as the dependent variable. Separate models were fitted for the mania–remission and depression–remission subsets.

The univariate mixed-effects model was specified as

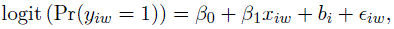

where *y_iw_* ∈ {0, 1} denotes the clinical state for week *w* of participant *i*, *x_iw_* represents the weekly aggregated actigraphic feature (either MEAN or VAR), *β*_0_ is the fixed intercept, and *β*_1_ is the fixed-effect coefficient representing the effect of the aggregated feature. The term *b_i_* ∼ N (0*, τ* ^2^) denotes a subject-specific random intercept capturing baseline differences between participants, and *ɛ_ij_* represents the residual, i.e., the portion of the response not explained by the systematic part of the model.

For research question Q3, multivariate mixed-effects models were fitted including both the weekly mean and the within-week standard deviation of each feature as regressors:

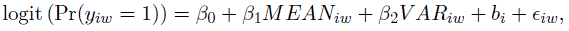

where *MEAN_iw_* denotes the weekly mean and *V AR_iw_* the within-week standard deviation of the given actigraphic feature. This model allowed us to assess whether variability is associated with the outcome beyond the mean level of the feature.

The models do not explicitly account for temporal autocorrelation within participants, which may lead to underestimated standard errors. However, the analysis was restricted to weekly observations with clearly defined clinical labels (remission, mania, or depression), while intermediate, mixed, or subthreshold states were excluded. This design choice reduces temporal ambiguity in the outcome definition and partially mitigates concerns regarding serial dependence, although it does not fully eliminate within-participant autocorrelation.

Random slopes were not included in the models because the limited number of manic or depressive relapse events per participant would not support reliable estimation of additional subject-specific slope effects.

All P values were adjusted for multiple comparisons using the Benjamini–Hochberg procedure to constrain the false discovery rate (FDR) to 5%, with significance for each comparison based on an adjusted P value <.05. In addition, the performance of all mixed-effects models was evaluated in terms of discriminative ability using the AUC. To ensure a population-level estimate of model performance, AUC values were derived from marginal probabilities based exclusively on fixed effects, thereby excluding subject-specific random intercepts and avoiding artificial inflation of discrimination estimates due to within-subject dependence.

### Analytical approach

To further assess the impact of data transformation on distributional properties, we examined the correlation coefficients between MEAN and VAR aggregates across the original and transformed datasets. This analysis was conducted to quantify changes in mean–variance coupling induced by the transformation procedure. Pearson correlation coefficients (*r*) (*r*) and AUCs were computed separately within the manic and depressive subsets to evaluate whether transformation differentially affected the structure of variability across mood states.

To assess whether variability effects reflected statistical artifacts induced by mean–variance coupling (Q1), we first estimated the association between the weekly mean of each feature and the clinical state using models fitted on the original data. Features for which both the MEAN and VAR regressors showed a statistically significant association were considered at risk for mean-driven VAR artifacts. We then examined the corresponding VAR models on transformed data. The comparison allowed us to determine whether variability remained associated with the outcome after accounting for mean–variance dependence.

To determine whether temporal variability distinguishes clinical state (Q2), we evaluated the VAR models fitted on the variance-stabilized (transformed) data. This analysis quantified the discriminative value of variability across all actigraphic features after suppressing the mean–variance coupling.

To assess whether variability provides additional information beyond the weekly mean (Q3), we compared nested mixed-effects logistic regression models using likelihood ratio tests. For each feature, a baseline model including only the transformed MEAN regressor was compared with an extended model including both the transformed MEAN and transformed VAR using likelihood ratio tests (LRT) and differences in Akaike information criterion (AIC). This allowed us to evaluate whether adding variability significantly improved model performance in distinguishing clinical states. Importantly, this analysis was restricted to features for which the transformed VAR regressors were statistically significant in prior steps (Q2), ensuring that model comparisons were conducted only on features with evidence of relevance and reducing the influence of noise or non-informative state markers.

The data flow diagram in Figure 1 summarizes all these parallel preprocessing steps and shows patient counts at each stage.

**Figure 1:**
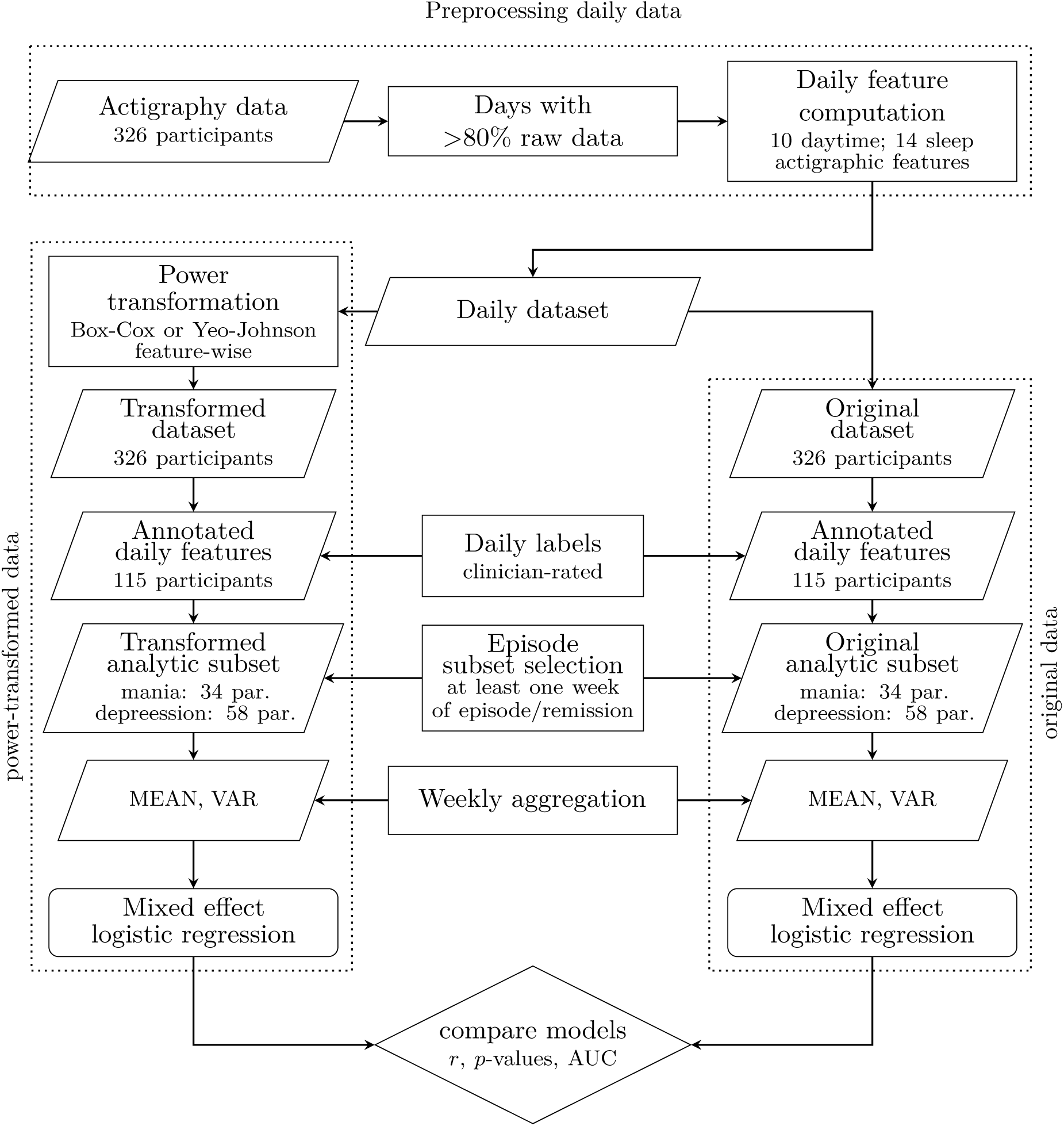
Parallel preprocessing and analysis pipelines for original and power-transformed actigraphic features. Actigraphy data from 326 participants were first filtered to retain days with more than 80% valid raw data, after which 10 daytime and 14 sleep actigraphic features were computed on a daily basis. The resulting daily feature dataset was analyzed using two parallel pipelines: (i) the original feature set and (ii) a feature-wise power-transformed feature set obtained using the Box–Cox or Yeo–Johnson transformation. Daily mood-state labels were automatically assigned to both feature sets using a classification algorithm that integrated clinician-administered Montgomery–Åsberg Depression Rating Scale (MADRS) and Young Mania Rating Scale (YMRS) scores, self-reported mood assessments (ASERT), and patients’ health records, yielding annotations for 115 participants. Participants were subsequently restricted to those with at least one week of remission and at least one week of a manic or depressive episode, resulting in separate analytic subsets for mania (34 participants) and depression (58 participants). Weekly feature aggregation (mean and variance) was performed independently within each pipeline, followed by fitting mixed-effects logistic regression models. Finally, the discriminative performance of models trained on the original and power-transformed features was compared using Pearson correlation coefficient (*r*), model significance (*p*-values), and the area under the receiver operating characteristic curve (AUC).

## Results

### Participants and Data

The initial cohort consisted of 326 participants with BD. All preprocessing steps requiring population-level parameter estimation, including variance-stabilizing transformations (Box–Cox or Yeo–Johnson), were performed on the full cohort before restricting the data to the analytical sample. Monthly MADRS and YMRS clinician ratings were available for 115 participants. Of these, 72 unique participants met the criteria for inclusion, having experienced at least one episode of mania or depression along with remission. This group formed two analytical subsets: 34 participants with mania–remission episodes (180 weeks of mania, 861 weeks of remission) and 58 participants with depression–remission episodes (490 weeks of depression, 1356 weeks of remission). The demographic, health, and activity characteristics of the participants included in each subset are summarized in Table 1.

**Table 1:** Demographic, health, and activity characteristics of participants for mixed-effects models.

|  | Mania subset * | Depression subset |
| --- | --- | --- |
| **Cohort overview** |  |  |
| Participants ** | 34 | 58 |
| Sex – female | 24 (71%) | 33 (57%) |
| Age | 35.0 ± 8.8 | 39.4 ± 10.7 |
| Body Mass Index (BMI) | 27.8 ± 6.1 | 29.6 ± 6.4 |
| **Actigraphy available days (per participant)** |  |  |
| Follow-up days | 955.6 ± 330.2 | 928.9 ± 357.9 |
| Mean percentage of valid days † | 81.3 ± 12.4 | 80.1 ± 14.0 |
| **Number of weekly ASERT questionnaires (individual counts)** |  |  |
| Number of questionnaires | 111.2 ± 41.5 | 103.3 ± 44.3 |
| Questionnaires per week | 1.0 ± 0.2 | 1.0 ± 0.2 |
| **Clinical scales (individual means)** |  |  |
| Baseline MADRS | 5.6 ± 5.7 | 8.1 ± 7.2 |
| Episode MADRS | 2.9 ± 2.7 | 20.4 ± 4.9 |
| Remission MADRS | 2.7 ± 1.5 | 3.4 ± 2.0 |
| Baseline YMRS | 4.0 ± 6.2 | 3.0 ± 4.5 |
| Episode YMRS | 15.5 ± 3.9 | 1.1 ± 1.4 |
| Remission YMRS | 2.0 ± 1.1 | 1.7 ± 1.3 |
| Episode intervention total ‡ | 25 | 32 |
| **Available episodes per participant (individual min / mean / max)** |  |  |
| Episode periods total | 1.0 / 3.0 / 9.0 | 1.0 / 3.8 / 13.0 |
| Remission periods total | 4.0 / 14.2 / 35.0 | 3 / 15.2 / 35.0 |
| Used episode weeks | 1.0 / 5.3 / 23.0 | 1.0 / 8.4 / 49.0 |
| Used remission weeks | 1.0 / 25.3 / 73.0 | 1.0 / 23.4 / 86.0 |
| **Total number of weeks used for regression model** |  |  |
| Total number of used episode weeks | 180 | 490 |
| Total number of used remission weeks | 861 | 1356 |

\* Includes both manic and hypomanic episodes as defined in the Definition of Clinical State and Data Annotation
\*\* Number of participants included in the analysis
† Days with at least 80% of available samples (also excludes non-wear periods)
‡ Defined as the sum of hospitalizations and sick leaves
Periods (= periods of continuous labeling, with no respect to the length of the episode)

### Power Transformation Methods

Figure 2 compares the distributions of actigraphic features between the original dataset and the datasets obtained after B-C and Y-J transformations.

**Figure 2:**
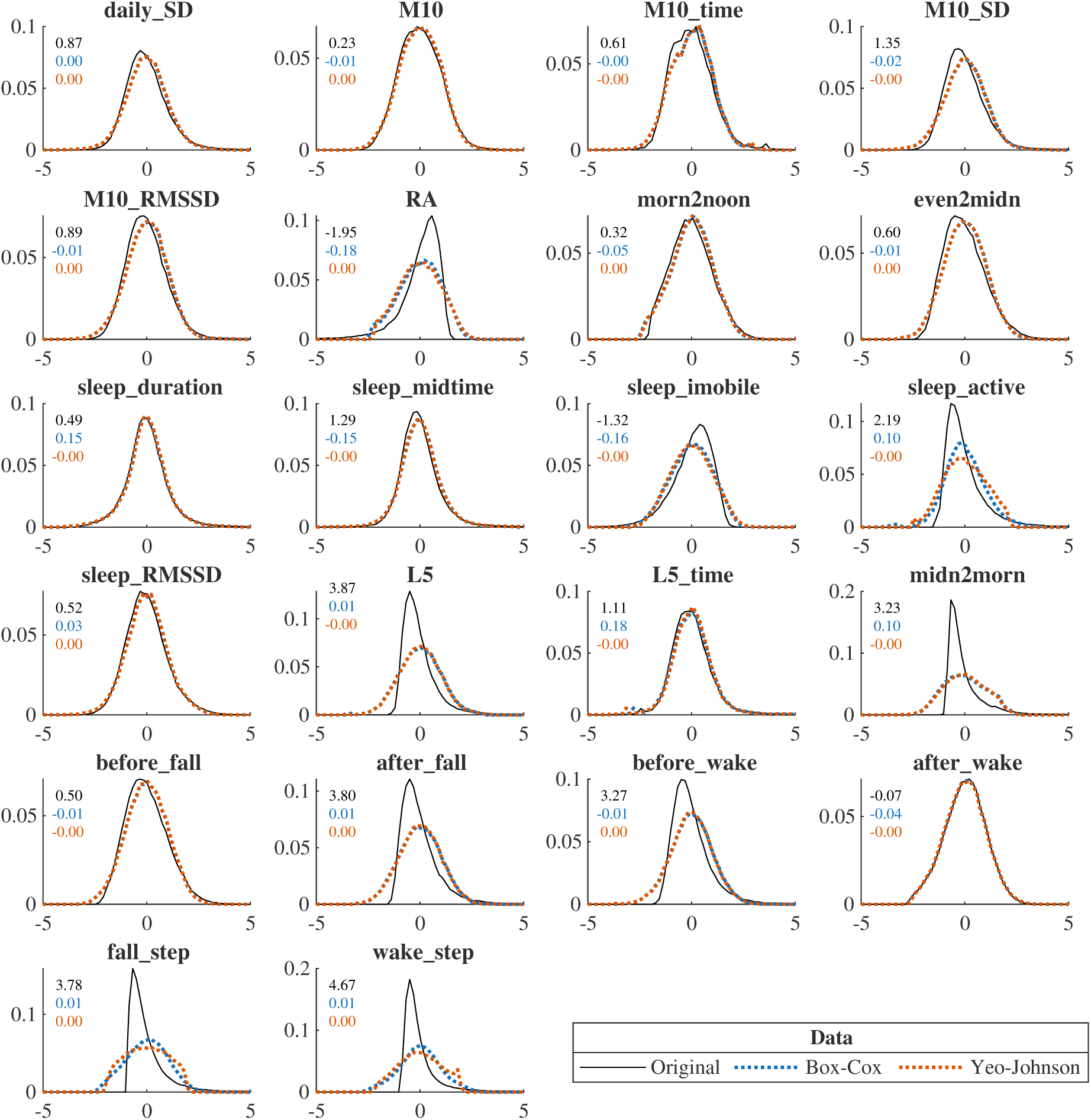
Visualizations of power transformation methods on actigraphy data. Standardized distributions of feature values in the original dataset (black) and after application of power transformations: Box–Cox (blue) and Yeo–Johnson (red), all based on the full study cohort of 326 participants. The transformations reduce the skewness of the data and, consequently, attenuate the dependence between the mean and the standard deviation while preserving the overall data structure. The skewness values of the original and transformed distributions are indicated in the figure using the corresponding color coding.

The power transformation, applied to the full study cohort, substantially reduced the correlation between MEAN and VAR aggregates. As illustrated in Figure 3, which provides a comparison with correlations computed on the original data, the mean absolute correlation decreased from 0.42 (0.27) to 0.06 (0.05), corresponding to an average reduction of 0.36 in the mania subset. Similarly, in the depressive subset, the mean absolute correlation declined from 0.43 (0.23) to 0.04 (0.04), representing an average reduction of 0.39. Full details are provided in Supplementary Table S1.

**Figure 3:**
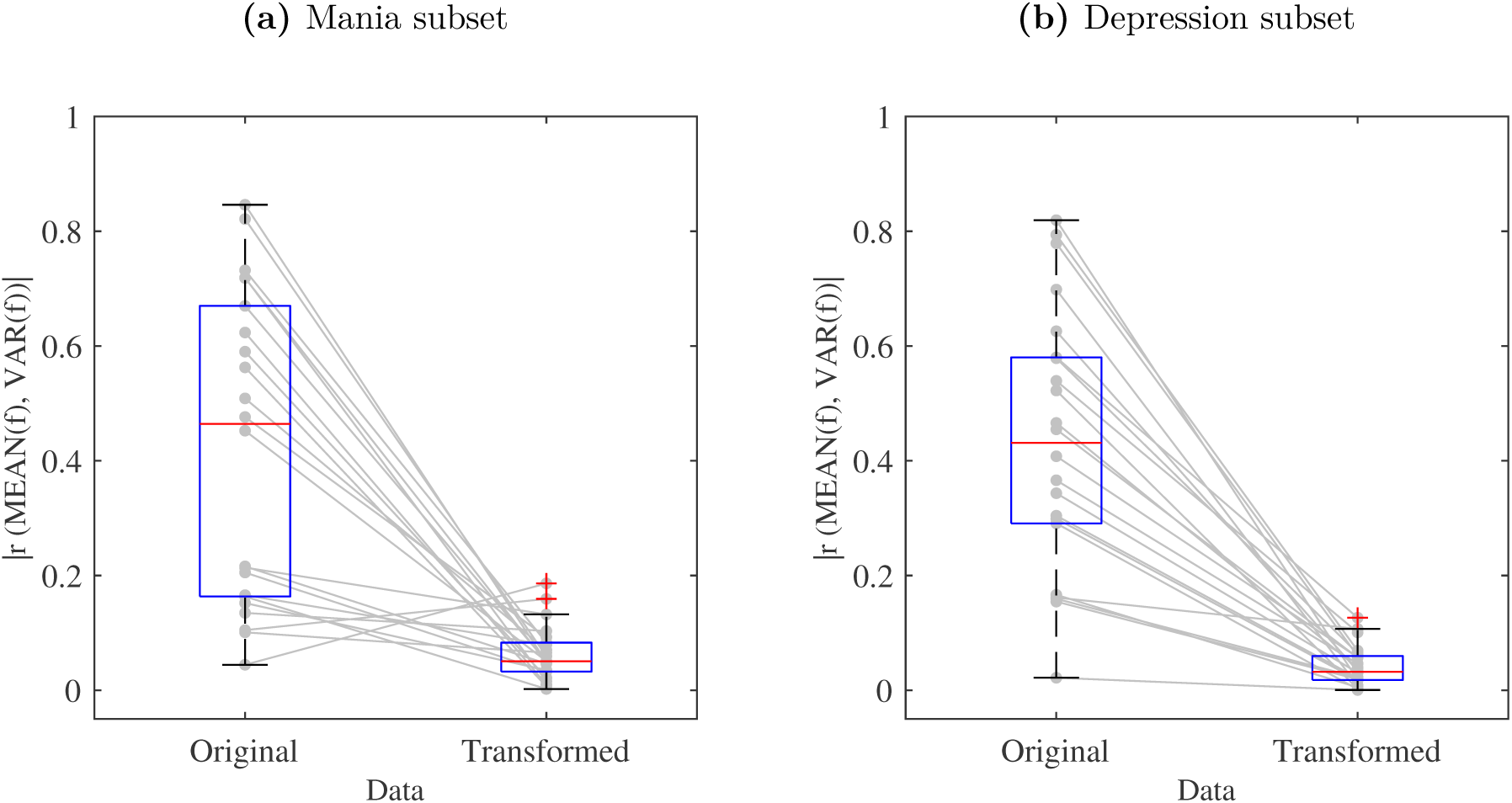
Suppression of mean–variability correlation across actigraphy features via power transformation. Boxplots of absolute Pearson correlation coefficients (*|r|*) between weekly MEAN and VAR aggregates before and after power transformation (Box-Cox or Yeo-Johnson), for each (n=22) actigraphic feature (f). **(a)** Mania subset; **(b)** Depression subset. The transformation consistently reduces the magnitude of correlation, indicating stabilization of the variance and weakening the statistical coupling between MEAN and VAR.

Furthermore, Figure 4 illustrates the effect of the variance-stabilizing transformation on the relationship between MEAN and VAR for a representative actigraphic feature—specifically, the mean activity level between 00:00 and 06:00 (midn2morn).

**Figure 4:**
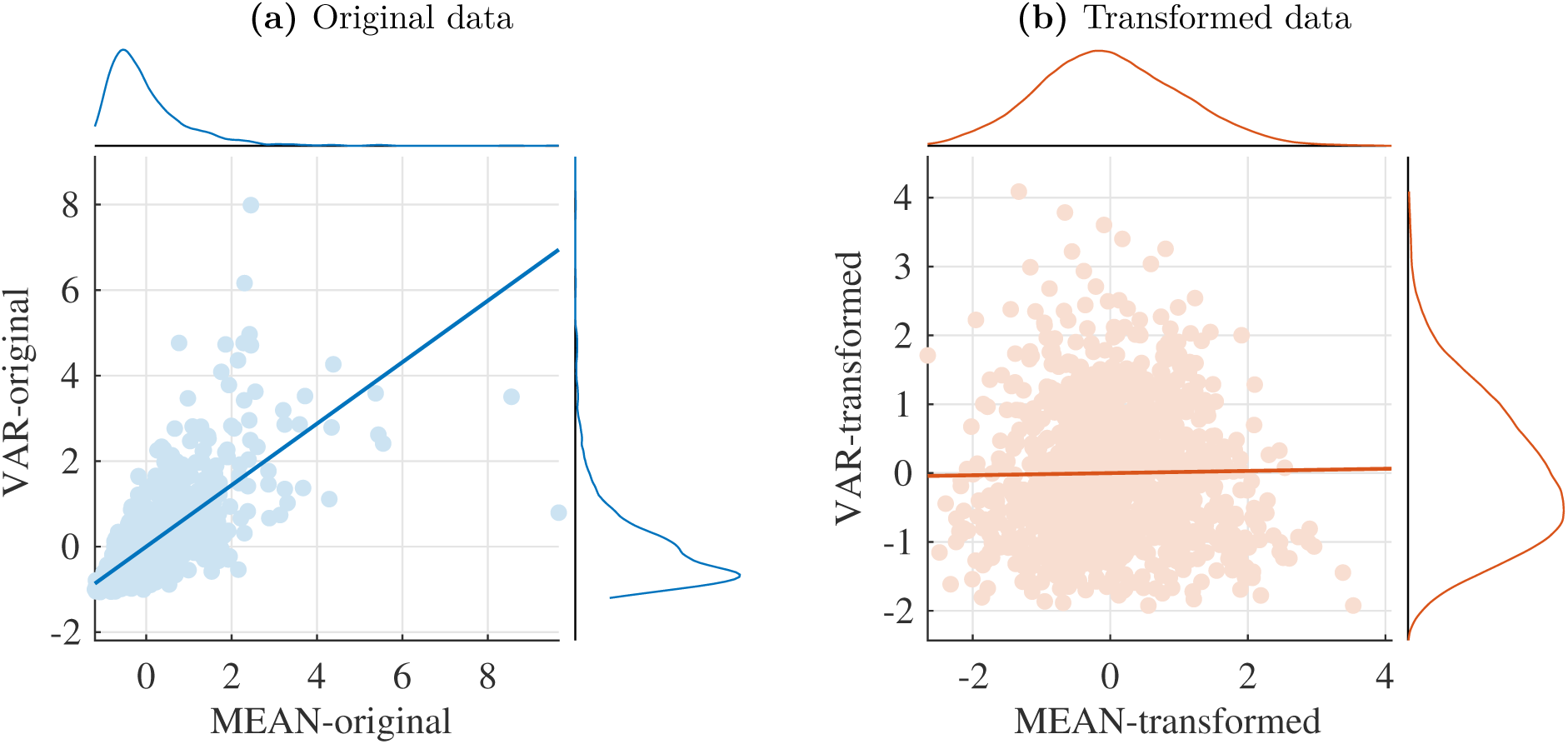
Example feature: Suppression of mean–variability correlation via power transformation. Scatter plots of standardized (z-score) original and power transformed weekly actigraphy aggregates—MEAN vs. VAR—before and after transformation, shown for a representative actigraphic feature: mean activity level between 00:00 am and 06:00 am (midn2morn), in the mania-remission subset. **(a)** Original data: strong positive correlation between MEAN and VAR; **(b)** Transformed data: reduced correlation after applying the Yeo-Johnson transformation. This decorrelation stabilizes the variance and weakens the statistical coupling between VAR and MEAN. As a result, the two summary statistics can be interpreted as less statistically dependent descriptors of the underlying distribution.

### Regression Modeling Results

The modelling analysis results for our three principal questions (Q1-Q3) are summarized in Table 2 and detailed below. For the individual P values, see the supplementary Table S2 (mania) and Table S3 (depression). For AUC values, see supplementary tables S4 (mania) and S5 (depression).

**Table 2:**
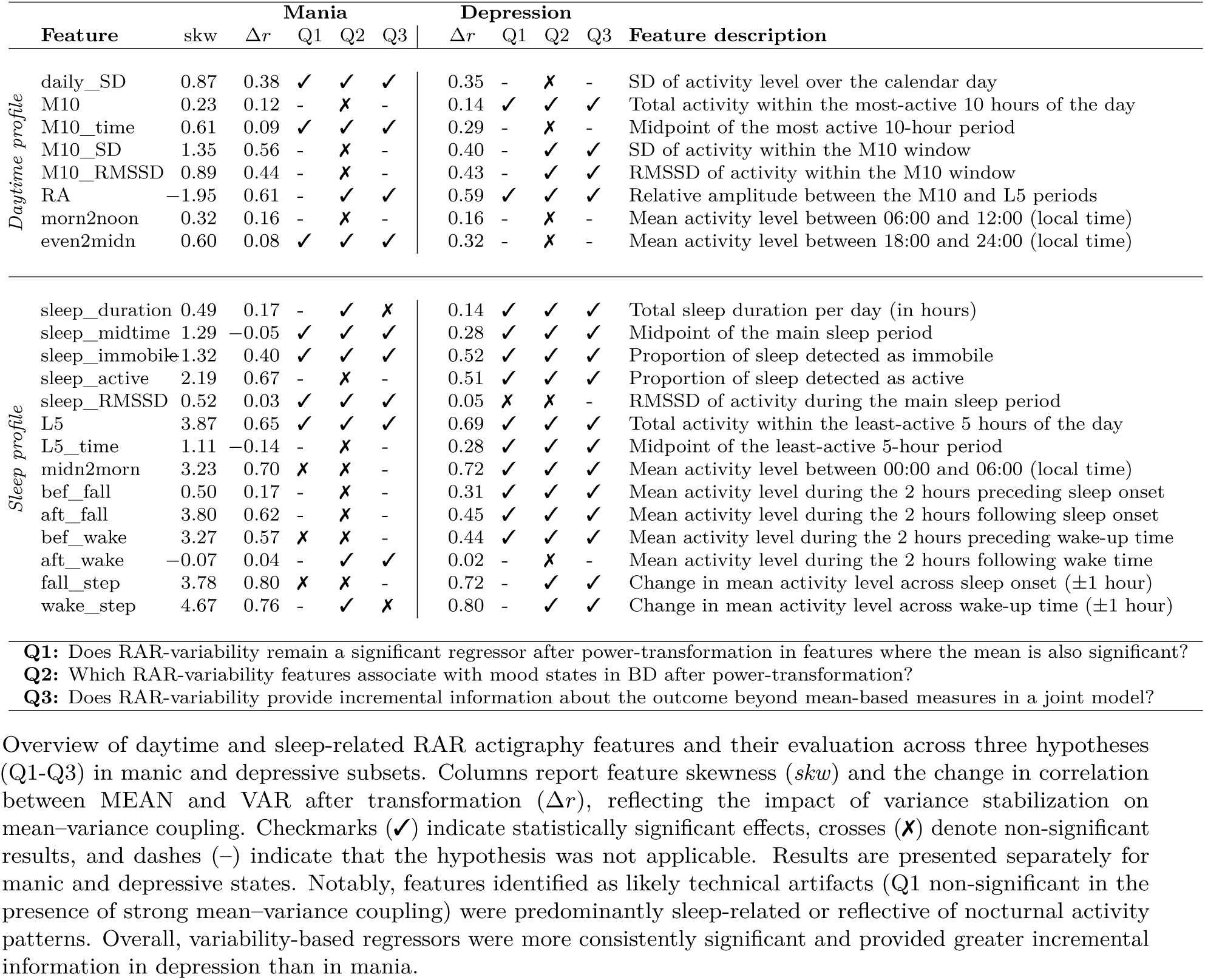
Actigraphic features and hypothesis testing results.

#### Q1: Is variability only a technical artifact?

To answer Q1, we focused on features at risk of spurious association with the clinical state, defined as features for which both the MEAN and VAR aggregates showed significant association with the outcome on the untransformed data. Then, we evaluated the association between the VAR regressor and the outcome on the transformed data. Here, the majority of variability regressors remained statistically significant after variance-stabilizing transformation, although effect sizes were modest. In the mania subset, 7 out of 10 features identified as potentially susceptible to mean–variance coupling remained significant after transformation, with model discrimination ranging from AUC = 0.54 to 0.63. In the depression subset, 12 out of 13 features remained significant, with AUC values between 0.51 and 0.58. These findings indicate that the effects of variability in most, though not all, actigraphy features are not solely attributable to mean–variance coupling. Associations with specific mood states remain significant even after controlling for the effect of the corresponding mean measures.

#### Q2: Is variability a standalone state marker?

In this analysis, the association between aggregates of all actigraphy features after the variance-stabilizing transformation were tested. The temporal variability showed significant associations with clinical state in both mania and depression subsets. In the mania subset, 11 out of 22 features were significant, distributed evenly across daily and sleep-based features, with AUC values ranging from 0.53 to 0.63. In the depression subset, 16 out of 22 features were significant: predominantly sleep-based features with AUC values between 0.50 and 0.58. Overall, variability regressors were more frequently significant in depression than in mania and focused on the sleep profile.

#### Q3: Does variability contribute information beyond the mean?

To see if variability carries additional outcome-relevant information on top of the mean, we focused on variability features that showed discriminative power on their own (Q2) and used a joint model, combining both MEAN and VAR regressors, and compared it to the MEAN-only model. The inclusion of VAR significantly improved model fit according to LRT for most features in both conditions (9/11 in mania and 16/16 in depression). However, these improvements translated into only modest increases in discriminative performance, with AUC values increasing from 0.50–0.65 to 0.54–0.66 in mania and from 0.49–0.57 to 0.52–0.59 in depression. These results support the results of Q2: the temporal variability represents a standalone biomarker in BD, even when controlling for the mean. Despite these improvements, gains in discrimination performance were modest.

## Discussion

This study examined whether temporal variability in actigraphy-derived RAR features constitutes a standalone state-marker in BD. Results from generalized mixed-effects models with a logistic link function indicate that temporal variability is indeed a standalone biomarker for most features, particularly for depression–remission discrimination, while mania–remission showed more instances of spurious associations driven by mean–variance coupling.

Variance-stabilizing transformations substantially reduced intra-feature MEAN–VAR correlations, enabling VAR-based features to act as decoupled regressors and improving their interpretability in multivariable models. While B-C suits strictly positive right-skewed variables, Y-J proved more broadly applicable, accommodating all skewness forms and negative values. All negatively skewed variables were sleep-related (relative amplitude, sleep immobility, post-wake activity), and Y-J was preferred for these. Sleep-related features showed the largest average reduction in correlation strength, indicating particular sensitivity to scale effects and distributional asymmetry. Following transformation, most variability features remained significant in the univariate models (7/10 in mania; 12/13 in depression). However, several sleep-linked features **(mania subset: mean activity between 00:00 and 06:00, activity before waking up, change in mean activity across sleep onset; depression subset: RMSSD of activity during main sleep)** lost significance after accounting for VAR collinearity, indicating their apparent discriminative value on the original scale was partly attributable to shared variance with global activity dispersion. Conversely, a small number of features **(mania subset: sleep duration, change in mean activity across wake-up time; depression subset: RMSSD of activity within the most-active 10 hours of the day, change in mean activity across wake-up time)** became significant only after transformation, suggesting that variance stabilization can unmask a genuine signal by reducing leverage from extreme observations and improving alignment with model assumptions.

In joint models, VAR inclusion improved discrimination beyond MEAN-only models in most cases (mean AUC gain: 4% in mania, 3% in depression; maximum: 12% and 7% respectively). Although statistically significant, these gains are unlikely to translate into substantial standalone clinical utility. Overall, feature significance and importance are conditional on scale, model specification, and collinearity structure: models on skewed untransformed data may overstate variability’s standalone contribution, while transformed data provide a more conservative and interpretable assessment.

### Clinical Interpretation

The results support treating variability in RAR actigraphy features as a clinically relevant signal in BD rather than a technical artifact, consistent with longitudinal evidence that amount, timing, and variability of rest–activity rhythms are among actigraphy measures most consistently associated with BD course [36], and with a recent review concluding that increased temporal variability may characterize BD [37]. This holds across temporal scales: [17] found that BD patients with unstable rest–activity cycles showed greater minute-to-minute variability despite comparable mean activity, while [12] demonstrated that complexity and variability metrics outperformed mean activity in distinguishing mood states, directly paralleling the present finding that transformed VAR was significant in 11/22 mania features and 16/22 depression features. The absolute discriminative performance (AUC: 0.50–0.66) is modest but consistent with the broader literature [2, 36].

Not all variability effects reflected independent clinical phenomena. In the mania subset, three nocturnal activity features **(mean activity between 00:00 and 06:00, activity before waking up, change in mean activity across sleep onset)** had significant VAR effects attributable to distributional confounding (skewness *>* 3.2; mean–variance correlation reduction *>* 0.5 after transformation). Their near-zero baseline during healthy sleep renders manic-episode elevations both clinically real and statistically extreme [14]. This artifact pattern was absent in the depression subset, where **RMSSD of activity during main sleep**, despite not reaching standalone significance (*P* = .083), still improved joint model fit, suggesting residual clinical information in its variability.

The persistence of variability effects after mean–variance decoupling supports the interpretation that variability encodes information not reducible to statistical dependency. [25] reported that variability/stability measures improved classification AUC from 0.64 to 0.82, a more pronounced gain than observed here, likely reflecting differences in sample characteristics and absence of explicit mean–variance decoupling. For depression specifically, [26] found that day-to-day step-count variability preceded depressive symptom onset by a median of 7 days, corroborating the present finding that depression VAR features were more robustly significant. Apparently divergent findings from [27], where reduced activity variability but increased sleep onset latency variability predicted depression, likely reflect differences in operational definitions and modalities, underscoring the need for feature-level analyses such as this one. For mania, [28] found that within-night sleep variability preceded hypomanic onset before activity changes emerged, consistent with the less consistent, more feature-dependent mania variability effects observed here. Early warning signal approaches further reinforce this picture: [13] demonstrated that variability dynamics predicted mood transitions up to four weeks in advance and outperformed mean-based indicators, suggesting the modest incremental gains in joint models here may translate into clinically meaningful lead times in prospective monitoring frameworks.

In joint models, the most significant univariate VAR regressors contributed incremental information beyond MEAN (9/11 in mania; 16/16 in depression), with the main exceptions being **sleep duration and wake-up time activity change in the mania subset**. The latter finding diverges from [30], who retained sleep duration variability in optimal models discriminating remitted BD from healthy controls, though that between-subject design is not directly comparable to the present within-patient episode–remission contrast. The stronger and more consistent incremental effects in depression than mania extend the pattern noted by [5], that sleep-related actigraphy measures show stronger associations in depression than mania, specifically to variability-based measures.

A notable finding was the complete non-overlap of significant variability regressors between mania–remission and depression–remission analyses across all three research questions, extending prior evidence of qualitatively distinct actigraphy profiles per polarity [15, 37]. This is further supported by [6], who found distinct circadian parameter profiles per polarity, and [8], whose inpatient findings converge with the present outpatient results. Mechanistically, depressive states appear to disrupt sleep timing and regularity, manifesting as increased variability in sleep midtime, L5 timing, and nocturnal patterns, whereas manic states disrupt activity thresholds and sleep architecture, producing distributional extremes more susceptible to mean–variance confounding. The closest precedent is [10], who found actigraphy discriminated depressive but not manic days from inter-episode states in BD outpatients, broadly consistent with the asymmetry observed here. Given differences in sample size and statistical power between subsets, the complete non-overlap should be interpreted cautiously and requires replication in larger cohorts.

Overall, variability in RAR features is not merely a statistical artifact: although mean–variance coupling can inflate variability effects, particularly in highly skewed features, most associations persisted after variance stabilization and provided complementary information beyond mean activity levels. The absolute discriminative performance was modest (AUC: 0.50–0.66), consistent with the exploratory design and reflecting constraints including limited subset sizes, strict state-labeling criteria, and the use of population-level rather than person-specific slopes. Crucially, high classification performance was not the primary objective, the analyses were designed to determine whether temporal variability carries information beyond the mean, not to optimize clinical prediction. Nevertheless, these findings strengthen the emerging evidence that variability is a clinically meaningful dimension of BD and demonstrate the importance of explicitly controlling for statistical artifacts when evaluating variability-based biomarkers.

### Limitations

The analysis has the following limitations, which have to be considered when interpreting the results:

In-sample regression performance does not provide evidence of out-of-sample generalizability; our primary aim was to investigate the impact of skewness rather than to optimize predictive performance.
Discretization into clinical episodes (mania, depression, remission) substantially reduces sample size and may introduce selection bias due to segmentation of continuous trajectories. On the other hand, it allows for a straightforward evaluation scenario in situations where the clinical labels depend on This limitation does not affect the primary technical objective of the study but should be considered when interpreting the results.
The effect of the one-week aggregation window on mean–variance coupling was not systematically evaluated. The seven-day window was selected to align with the inherent weekly periodicity of the actigraphy data. However, temporal aggregation may affect both variance structure and dependence estimates.

## Conclusion

This study addressed three key questions regarding variability in RAR actigraphy features in BD.

First (Q1), variability in RAR biomarkers cannot be considered a mere technical artifact. Although mean–variance coupling substantially contributed to some associations, particularly in highly skewed nocturnal features in the mania subset, variance-stabilizing transformations markedly reduced this dependency, and the majority of variability effects persisted (7 of 10 in mania; 12 of 13 in depression), indicating that they are not fully explained by statistical coupling. Features identified as likely artifacts were predominantly sleep-related or captured nocturnal activity patterns, consistent with the distinct activity dynamics known to characterize manic states.

Second (Q2), variability remained a significant state marker of clinically relevant mood states in the majority of features after adjusting for mean influence (11 of 22 in mania; 16 of 22 in depression), supporting the interpretation that variability captures meaningful aspects of behavioral and circadian dysregulation in BD that are not reducible to central tendency.

Third (Q3), variability provided incremental outcome-relevant information beyond mean-based measures in most evaluated features, with joint MEAN+VAR models outperforming MEAN-only models across both subsets. Discrimination gains were modest (3–4% on average; up to 12% in mania and 7% in depression), suggesting that mean and variability reflect complementary dimensions of the underlying processes, while standalone clinical utility remains limited.

Overall, significant variability-based state markers were entirely non-overlapping between manic and depressive subsets across all three research questions, extending prior evidence of polarity-specific actigraphy signatures to variability features tested with explicit artifact control. Variability showed stronger and more consistent performance in the depression subset, possibly because the seven-day observation window captures depressive dynamics more effectively than manic ones. While standalone clinical utility is limited, variability measures represent a non-redundant and informative component for modeling mood states in BD, particularly when appropriately transformed. Variance-stabilizing transformations should be considered a standard preprocessing step in actigraphy research, and future work should evaluate variability within multivariate and multimodal frameworks to fully realise its potential as a monitoring biomarker.

## Acknowledgments

Authors CAK, EB, and JS were responsible for the original study conception and design. CAK performed data analyses and related calculations. JS was responsible for preprocessing the raw data and computing daily actigraphic features. EB and JS were responsible for the scientific supervision of the study and provided regular guidance and feedback throughout the research process. MK, FS, and JS were responsible for data collection. CAK wrote the first draft of the manuscript. CAK, EB, JS, MK, MA, and FS were responsible for the interpretation of the results. All authors performed critical revision of the manuscript and approved the final version.

This work was supported by the Brain Dynamics project, funded by the Operational Programme Johannes Amos Comenius, no. CZ.02.01.01/00/22_008/0004643, and by the Ministry of Health of the Czech Republic, grant no. NU23-04-00534.

## Data Availability

The datasets used during the current study are available from the corresponding author on reasonable request.

## Multimedia Appendix 1

Supplementary tables for the power transformation results (feature-wise skewnesses, selected transformations, Pearson correlation coefficients), and performance matrices for MEAN, VAR, and joint models (*p*-values, LRT, Δ AIC, AUC).

## Abbreviations

*r*: Pearson correlation coefficient. 7
AIC: Akaike information criterion. 7, 17
ASERT: Aktibipo Self-rating Ecological Momentary Assessment. 4
AUC: area under the receiver operating characteristic curve. 3, 7, 12, 13, 15, 17
B-C: Box-Cox power-transformation. 5, 6, 9, 13
BD: bipolar disorder. 2–4, 6, 9, 13–16
GLMM: generalized linear mixed-effects model. 6
LRT: likelihood ratio tests. 7, 13, 17
MADRS: Montgomery-Åsberg Depression Rating Scale. 4, 9
MEAN: mean-based approach. 5–8, 10–14, 16, 17, 21–24
RAR: rest-activity-rhythm. 2–4, 6, 12–15
VAR: time-variability-based approach. 5–8, 10–14, 16, 17, 21–24
Y-J: Yeo-Johnson power-transformation. 5, 9, 13
YMRS: Young Mania Rating Scale. 4, 9

## Supplementary Material

**Table S1:** Power transformation results.

|  | Feature | skw | Mania subset |  |  |  | Depression subset |  |  |  |
| --- | --- | --- | --- | --- | --- | --- | --- | --- | --- | --- |
| | | | T | $r_{orig}$ | $r_{trans}$ | $\Delta r$ | T | $r_{orig}$ | $r_{trans}$ | $\Delta r$ |
| Daytime profile | daily_SD | 0.87 | B-C | 0.48 | 0.09 | 0.38 | B-C | 0.41 | 0.06 | 0.35 |
|  | M10 | 0.23 | B-C | 0.15 | 0.04 | 0.12 | B-C | 0.15 | 0.02 | 0.14 |
|  | M10_time | 0.61 | Y-J | 0.17 | -0.08 | 0.09 | Y-J | 0.29 | 0.00 | 0.29 |
|  | M10_SD | 1.35 | B-C | 0.56 | -0.01 | 0.56 | Y-J | 0.45 | -0.06 | 0.40 |
|  | M10_RMSSD | 0.89 | B-C | 0.51 | 0.07 | 0.44 | B-C | 0.47 | 0.04 | 0.43 |
|  | RA | -1.95 | Y-J | -0.62 | 0.01 | 0.61 | Y-J | -0.63 | -0.03 | 0.59 |
|  | morn2noon | 0.32 | Y-J | 0.16 | 0.00 | 0.16 | Y-J | 0.17 | -0.01 | 0.16 |
|  | even2midn | 0.60 | Y-J | 0.21 | -0.13 | 0.08 | Y-J | 0.37 | 0.05 | 0.32 |
| Sleep profile | sleep_duration | 0.49 | Y-J | 0.21 | 0.03 | 0.17 | B-C | 0.16 | -0.02 | 0.14 |
|  | sleep_midtime | 1.29 | Y-J | 0.10 | -0.16 | -0.05 | Y-J | 0.30 | -0.02 | 0.28 |
|  | sleep_immobile | -1.32 | Y-J | -0.45 | 0.05 | 0.40 | Y-J | -0.52 | 0.00 | 0.52 |
|  | sleep_active | 2.19 | Y-J | 0.72 | -0.05 | 0.67 | Y-J | 0.58 | -0.06 | 0.51 |
|  | sleep_RMSSD | 0.52 | B-C | 0.13 | -0.10 | 0.03 | B-C | 0.16 | -0.11 | 0.05 |
|  | L5 | 3.87 | B-C | 0.73 | 0.08 | 0.65 | Y-J | 0.70 | 0.01 | 0.69 |
|  | L5_time | 1.11 | B-C | 0.04 | -0.19 | -0.14 | B-C | 0.30 | -0.02 | 0.28 |
|  | midn2morn | 3.23 | Y-J | 0.72 | 0.02 | 0.70 | Y-J | 0.78 | -0.06 | 0.72 |
|  | bef_fall | 0.50 | Y-J | 0.22 | -0.05 | 0.17 | Y-J | 0.34 | 0.03 | 0.31 |
|  | aft_fall | 3.80 | Y-J | 0.67 | 0.05 | 0.62 | Y-J | 0.58 | -0.13 | 0.45 |
|  | bef_wake | 3.27 | Y-J | 0.59 | -0.02 | 0.57 | Y-J | 0.54 | -0.10 | 0.44 |
|  | aft_wake | -0.07 | Y-J | -0.10 | -0.07 | 0.04 | Y-J | -0.02 | 0.00 | 0.02 |
|  | fall_step | 3.78 | B-C | 0.85 | 0.05 | 0.80 | B-C | 0.79 | -0.07 | 0.72 |
|  | wake_step | 4.67 | Y-J | 0.82 | -0.06 | 0.76 | Y-J | 0.82 | 0.02 | 0.80 |

Feature-wise skewness (skw) and selected variance-stabilizing transformation (T; Box-Cox or Yeo-Johnson). The columns $r_{orig}$ and $r_{trans}$ report Pearson correlations between weekly mean and variability aggregates before and after transformation, respectively. $\Delta r$ quantifies the absolute attenuation of mean-variance coupling following transformation. Results are presented separately for the mania and depression subsets.

**Table S2:** Mania-remission subset: Comparison of model P values and information criterion.

|  | Feature | Original data |  | Transformed data |  |  |  |
| --- | --- | --- | --- | --- | --- | --- | --- |
| | | MEAN | VAR | MEAN | VAR | LRT | $\Delta$ AIC |
| *Daytime profile* | daily_SD | <.001 | <.001 | <.001 | <.001 | <.001 | −20.49 |
|  | M10 | <.001 | .209 | <.01 | .456 | .619 | 1.66 |
|  | M10_time | <.05 | <.001 | <.01 | <.001 | <.001 | −48.45 |
|  | M10_SD | .150 | .158 | .199 | .350 | .289 | 0.34 |
|  | M10_RMSSD | .870 | .178 | .532 | .128 | .127 | −1.05 |
|  | RA | .210 | <.001 | .531 | <.001 | <.001 | −38.32 |
|  | morn2noon | <.001 | .802 | <.001 | .456 | .480 | 1.27 |
|  | even2midn | <.001 | <.001 | <.001 | <.001 | <.001 | −67.35 |
| *Sleep profile* | sleep_duration | <.001 | .159 | <.001 | <.01 | .063 | −2.76 |
|  | sleep_midtime | <.001 | <.05 | <.001 | <.01 | <.01 | −6.69 |
|  | sleep_immobile | <.05 | <.001 | <.01 | <.001 | <.001 | −48.33 |
|  | sleep_active | .973 | .593 | .385 | .630 | .688 | 1.80 |
|  | sleep_RMSSD | <.001 | <.001 | <.001 | <.001 | <.001 | −18.08 |
|  | L5 | <.01 | <.001 | .225 | <.001 | <.001 | −77.99 |
|  | L5_time | .077 | .415 | <.05 | .575 | .694 | 1.85 |
|  | midn2morn | <.001 | <.001 | <.001 | .294 | .293 | 0.45 |
|  | bef_fall | <.01 | .097 | <.01 | .456 | .619 | 1.64 |
|  | aft_fall | <.001 | .294 | <.001 | .206 | .087 | −1.90 |
|  | bef_wake | <.001 | <.001 | <.001 | .121 | .087 | −1.80 |
|  | aft_wake | .906 | <.01 | .887 | <.01 | <.01 | −8.79 |
|  | fall_step | <.001 | <.001 | <.001 | .227 | .426 | 1.05 |
|  | wake_step | <.001 | .691 | <.001 | <.01 | .063 | −2.62 |

P values of the MEAN and the VAR models estimated using the original and transformed data. The column LRT reports P values from a likelihood ratio test comparing the MEAN-only model with the joint model including both MEAN and VAR. $\Delta$ AIC denotes the change in the Akaike information criterion after inclusion of the VAR component.

**Table S3:** Depression-remission subset: Comparison of model P values and information criterion.

|  | Feature | Original data |  | Transformed data |  |  |  |
| --- | --- | --- | --- | --- | --- | --- | --- |
| | | MEAN | VAR | MEAN | VAR | LRT | $\Delta$ AIC |
| *Daytime profile* | daily_SD | <.05 | .225 | .054 | .410 | .499 | 1.54 |
|  | M10 | <.001 | <.05 | <.001 | <.001 | <.01 | −7.47 |
|  | M10_time | <.01 | .134 | <.05 | .158 | .121 | −0.63 |
|  | M10_SD | .318 | <.001 | .792 | <.001 | <.001 | −17.78 |
|  | M10_RMSSD | .054 | .402 | <.05 | <.001 | <.01 | −10.17 |
|  | RA | <.001 | <.001 | <.001 | <.001 | <.001 | −32.14 |
|  | morn2noon | <.001 | .926 | <.001 | .331 | .467 | 1.42 |
|  | even2midn | <.01 | .559 | <.01 | .317 | .423 | 1.24 |
| *Sleep profile* | sleep_duration | <.001 | <.001 | <.001 | <.001 | <.001 | −46.34 |
|  | sleep_midtime | <.001 | <.001 | <.001 | <.001 | <.001 | −14.59 |
|  | sleep_immobile | <.001 | <.01 | <.001 | <.01 | <.01 | −8.99 |
|  | sleep_active | <.001 | <.001 | <.001 | <.01 | <.001 | −16.23 |
|  | sleep_RMSSD | <.001 | <.01 | <.001 | .083 | <.05 | −3.94 |
|  | L5 | <.01 | <.001 | .955 | <.001 | <.001 | −39.08 |
|  | L5_time | <.001 | <.001 | <.001 | <.01 | <.01 | −8.00 |
|  | midn2morn | <.001 | <.001 | <.001 | <.01 | <.01 | −7.74 |
|  | bef_fall | <.05 | <.01 | .065 | <.01 | <.01 | −5.68 |
|  | aft_fall | <.001 | <.001 | <.01 | <.01 | <.001 | −13.15 |
|  | bef_wake | <.05 | <.001 | .245 | <.001 | <.001 | −19.88 |
|  | aft_wake | <.05 | .094 | <.05 | .083 | .083 | −1.33 |
|  | fall_step | .164 | <.001 | <.05 | <.001 | <.001 | −70.78 |
|  | wake_step | .051 | .203 | .532 | <.05 | <.05 | −4.22 |

P values of the MEAN and the VAR models estimated using the original and transformed data. The column LRT reports P values from a likelihood ratio test comparing the MEAN-only model with the joint model including both MEAN and VAR. $\Delta$ AIC denotes the change in the Akaike information criterion after inclusion of the VAR component.

**Table S4:** Mania-remission subset: AUC.

|  |  | Original data |  | Transformed data |  |  |
| --- | --- | --- | --- | --- | --- | --- |
| Feature |  | MEAN | VAR | MEAN | VAR | MEAN+VAR |
| *Daytime profile* | daily_SD | 0.58 | 0.57 | 0.57 | 0.55 | 0.59 |
|  | M10 | 0.54 | 0.48 | 0.54 | 0.48 | 0.54 |
|  | M10_time | 0.53 | 0.58 | 0.54 | 0.60 | 0.60 |
|  | M10_SD | 0.51 | 0.48 | 0.51 | 0.49 | 0.52 |
|  | M10_RMSSD | 0.51 | 0.49 | 0.51 | 0.48 | 0.49 |
|  | RA | 0.49 | 0.57 | 0.52 | 0.59 | 0.59 |
|  | morn2noon | 0.61 | 0.51 | 0.61 | 0.50 | 0.61 |
|  | even2midn | 0.56 | 0.60 | 0.55 | 0.58 | 0.61 |
| *Sleep profile* | sleep_duration | 0.64 | 0.52 | 0.65 | 0.53 | 0.66 |
|  | sleep_midtime | 0.57 | 0.53 | 0.57 | 0.54 | 0.58 |
|  | sleep_immobile | 0.55 | 0.55 | 0.56 | 0.58 | 0.60 |
|  | sleep_active | 0.47 | 0.51 | 0.53 | 0.52 | 0.52 |
|  | sleep_RMSSD | 0.54 | 0.56 | 0.54 | 0.58 | 0.58 |
|  | L5 | 0.53 | 0.60 | 0.51 | 0.63 | 0.63 |
|  | L5_time | 0.53 | 0.46 | 0.53 | 0.47 | 0.52 |
|  | midn2morn | 0.59 | 0.59 | 0.59 | 0.52 | 0.60 |
|  | bef_fall | 0.53 | 0.51 | 0.53 | 0.50 | 0.53 |
|  | aft_fall | 0.55 | 0.51 | 0.55 | 0.53 | 0.56 |
|  | bef_wake | 0.55 | 0.54 | 0.55 | 0.52 | 0.55 |
|  | aft_wake | 0.50 | 0.54 | 0.50 | 0.54 | 0.54 |
|  | fall_step | 0.56 | 0.56 | 0.55 | 0.52 | 0.56 |
|  | wake_step | 0.53 | 0.51 | 0.56 | 0.55 | 0.58 |

Discriminative performance of the mixed-effects logistic regression models. The table reports the area under the receiver operating characteristic curve (AUC) for each model. AUC values were calculated using marginal probabilities (fixed effects only) to provide a population-level estimate of model discrimination, excluding participant-specific random intercepts.

**Table S5:**
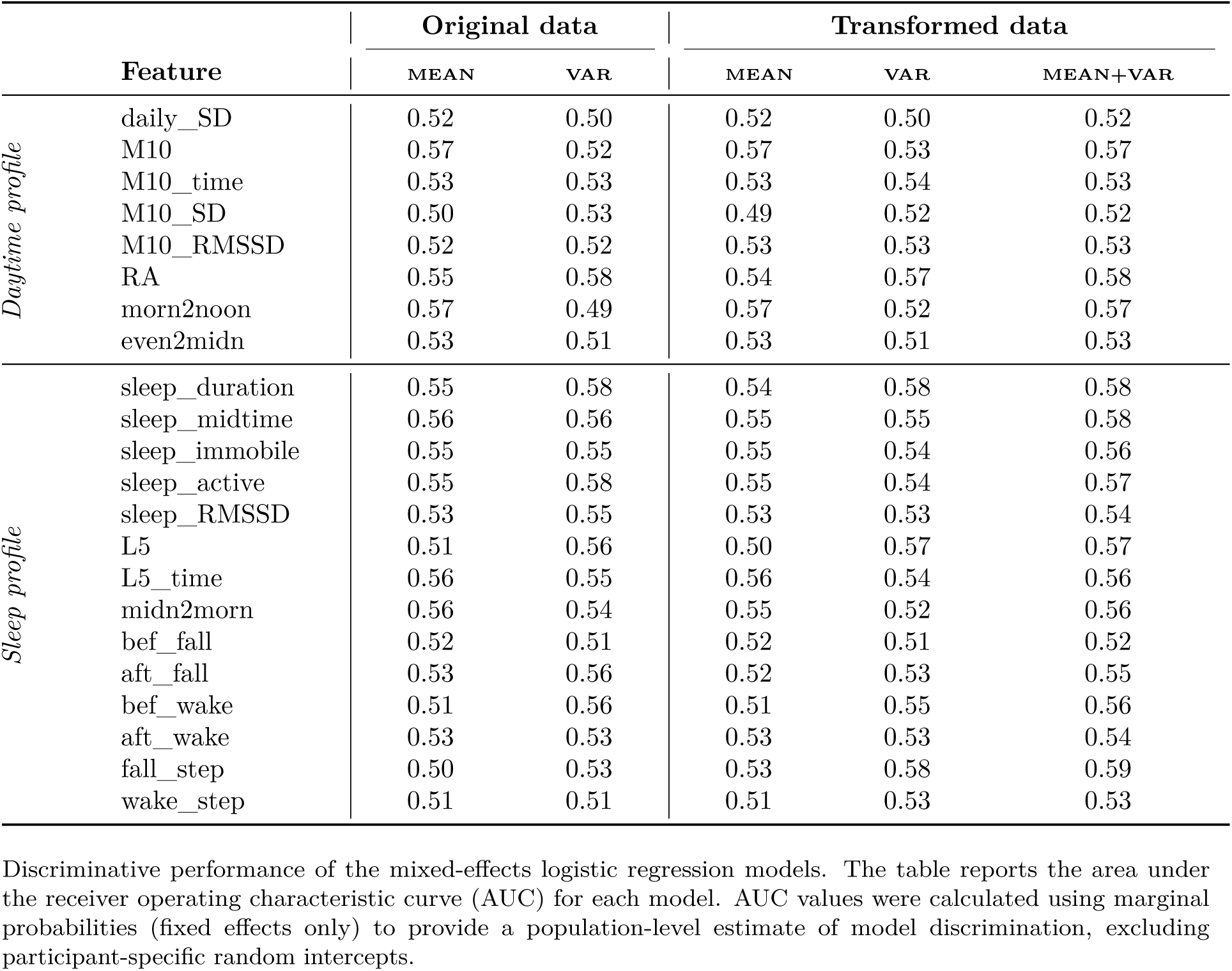
Depression-remission subset: AUC.

## References

[1] Scott J, Murray G, Henry C, Morken G, Scott E, Angst J, et al. Activation in bipolar disorders a systematic review. JAMA Psychiatry. 2017 2;74:189–96. PMID: 28002572.

[2] Tazawa Y, Wada M, Mitsukura Y, Takamiya A, Kitazawa M, Yoshimura M, et al. Actigraphy for evaluation of mood disorders: A systematic review and meta-analysis. Journal of Affective Disorders. 2019 6;253:257–69. PMID: 31060012.

[3] Martin JL, Hakim AD. Wrist actigraphy. Chest. 2011 6;139:1514-27. PMID: 21652563.

[4] Melo MCA, Abreu RLC, Neto VBL, de Bruin PFC, de Bruin VMS. Chronotype and circadian rhythm in bipolar disorder: A systematic review. Sleep Medicine Reviews. 2017 8;34:46–58. PMID: 27524206.

[5] Crescenzo FD, Economou A, Sharpley AL, Gormez A, Quested DJ. Actigraphic features of bipolar disorder: A systematic review and meta-analysis. Sleep medicine reviews. 2017 6;33:58–69. PMID: 28185811.

[6] Clemens J, Mühlbauer E, Reinhard I, et al. Circadian rhythm parameters differentiate euthymic, manic and depressive mood states in bipolar disorders – an explorative pilot study. International Journal of Bipolar Disorders. 2025;13:30. PMID: 41145890.

[7] Faurholt-Jepsen M, Brage S, Vinberg M, Kessing LV. State-related differences in the level of psychomotor activity in patients with bipolar disorder – Continuous heart rate and movement monitoring. Psychiatry Research. 2016;237:166–74. PMID: 26832835.

[8] Zhang Y, Deng X, Wang X, Luo H, Lei X, Luo Q. Can daily actigraphic profiles distinguish between different mood states in inpatients with bipolar disorder? An observational study. Frontiers in Psychiatry. 2023;14. PMID: 37363166.

[9] Wüthrich F, Nabb CB, Mittal VA, Shankman SA, Walther S. Actigraphically measured psychomotor slowing in depression: systematic review and meta-analysis. Psychological Medicine. 2022;52(7):1208–21. PMID: 35550677.

[10] Gershon A, Ram N, Johnson SL, Harvey AG, Zeitzer JM. Daily actigraphy profiles distinguish depressive and interepisode states in bipolar disorder. Clinical Psychological Science. 2016 7;4:641–50. PMID: 27642544.

[11] Salvatore P, Ghidini S, Zita G, Panfilis CD, Lambertino S, Maggini C, et al. Circadian activity rhythm abnormalities in ill and recovered bipolar I disorder patients. Bipolar Disorders. 2008 3;10:256–65. PMID: 18271904.

[12] Jakobsen P, Stautland A, Riegler MA, Côté-Allard U, Sepasdar Z, Nordgreen T, et al. Complexity and variability analyses of motor activity distinguish mood states in bipolar disorder. PLoS ONE. 2022 5;17. PMID: 35061801.

[13] Kunkels YK, Riese H, Knapen SE, et al. Efficacy of early warning signals and spectral periodicity for predicting transitions in bipolar patients: An actigraphy study. Translational Psychiatry. 2021;11(1):350. PMID: 34099627.

[14] Gonzalez R, Tamminga CA, Tohen M, Suppes T. The relationship between affective state and the rhythmicity of activity in bipolar disorder. Journal of Clinical Psychiatry. 2014;75. PMID: 24500063.

[15] Krane-Gartiser K, Henriksen TEG, Morken G, Vaaler A, Fasmer OB. Actigraphic assessment of motor activity in acutely admitted inpatients with bipolar disorder. PLoS ONE. 2014;9. PMID: 24586883.

[16] Cuesta-Frau D, Schneider J, Bakstein E, Vostatek P, Spaniel F, Novak D. Classification of Actigraphy Records from Bipolar Disorder Patients Using Slope Entropy: A Feasibility Study. Entropy. 2020;22(11):1243. PMID: 33287011.

[17] Krane-Gartiser K, Steinan MK, Langsrud K, Vestvik V, Sand T, Fasmer OB, et al. Mood and motor activity in euthymic bipolar disorder with sleep disturbance. Journal of Affective Disorders. 2016;202:23–31. PMID: 27253213.

[18] Song YM, Jeong J, de Los Reyes AA, Lim D, Cho CH, Yeom JW, et al. Causal dynamics of sleep, circadian rhythm, and mood symptoms in patients with major depression and bipolar disorder: insights from longitudinal wearable device data. eBioMedicine. 2024;103:105094. PMID: 38579366.

[19] Lipschitz JM, Lin S, Saghafian S, Pike CK, Burdick KE. Digital phenotyping in bipolar disorder: Using longitudinal Fitbit data and personalized machine learning to predict mood symptomatology. Acta Psychiatrica Scandinavica. 2025;151(3):434–47. PMID: 39397313.

[20] Lim D, Jeong J, Song YM, Cho CH, Yeom JW, Lee T, et al. Accurately predicting mood episodes in mood disorder patients using wearable sleep and circadian rhythm features. npj Digital Medicine. 2024;7:324. PMID: 39557997.

[21] Wu CT, Hsieh MH, Chen IM, Jhao LY, Liu DS, Wang SM, et al. Using Wearable Device and Machine Learning to Predict Mood Symptoms in Bipolar Disorder: Development and Usability Study. JMIR Medical Informatics. 2025;13:e66277. PMID: 40957006.

[22] Cho CH, Lee T, Kim MG, In HP, Kim L, Lee HJ. Mood Prediction of Patients With Mood Disorders by Machine Learning Using Passive Digital Phenotypes Based on the Circadian Rhythm: Prospective Observational Cohort Study. Journal of Medical Internet Research. 2019;21(4):e11029. PMID: 30994461.

[23] Schneider J, Bakštein E, Kolenič M, Vostatek P, Correll CU, Novák D, et al. Motor activity patterns can distinguish between interepisode bipolar disorder patients and healthy controls. CNS Spectrums. 2022;27:82–92. PMID: 32883376.

[24] Jakobsen P, Côté-Allard U, Riegler MA, Stabell LA, Stautland A, Nordgreen T, et al. Early warning signals observed in motor activity preceding mood state change in bipolar disorder. Bipolar Disorders. 2024;26(5):468–78. PMID: 38639725.

[25] Ferrand L, Hennion V, Godin O, Bellivier F, Scott J, Etain B. Which Actigraphy Dimensions Predict Longitudinal Outcomes in Bipolar Disorders? Journal of Clinical Medicine. 2022 4;11. PMID: 35456294.

[26] Ortiz A, Halabi R, Alda M, DeShaw A, Husain MI, Nunes A, et al. Day-to-day variability in activity levels detects transitions to depressive symptoms in bipolar disorder earlier than changes in sleep and mood. International Journal of Bipolar Disorders. 2025;13(1):13. PMID: 40175826.

[27] Halabi R, Mulsant BH, Tolend M, Blumberger DM, DeShaw A, Hintze A, et al. A systematic exploration of digital biomarkers for the detection of depressive episodes in bipolar disorder. npj Mental Health Research. 2026;5(1):13. PMID: 41724810.

[28] Ortiz A, Halabi R, Alda, Burgos A, DeShaw A, Gonzalez-Torres C, et al. Day-to-day variability in sleep and activity predict the onset of a hypomanic episode in patients with bipolar disorder. Journal of Affective Disorders. 2025;374:75–83. PMID: 39793618.

[29] Faedda GL, Ohashi K, Hernandez M, McGreenery CE, Grant MC, Baroni A, et al. Actigraph measures discriminate pediatric bipolar disorder from attention-deficit/hyperactivity disorder and typically developing controls. J Child Psychol Psychiatry. 2016 Jun;57(6):706–16. PMID: 26799153.

[30] Geoffroy PA, Boudebesse C, Bellivier F, Lajnef M, Henry C, Leboyer M, et al. Sleep in remitted bipolar disorder: A naturalistic case-control study using actigraphy. Journal of Affective Disorders. 2014;158:1–7. PMID: 24655758. Available from: 10.1016/j.jad.2014.01.012.

[31] Anýž J, Bakštein E, Dally A, Kolenič M, Hlinka J, Hartmannová T, et al. Validity of the aktibipo self-rating questionnaire for the digital self-assessment of mood and relapse detection in patients with bipolar disorder: Instrument validation study. JMIR Mental Health. 2021;8:1–17. PMID: 34383689.

[32] Lynch I, Harmata GIS, Barsotti EJ, Fiedorowicz JG, Williams AJ, Linkenmeyer C, et al. Feasibility and accuracy of the ASERT digital questionnaire in mood tracking for a longitudinal research study on bipolar disorder. Journal of Mood & Anxiety Disorders. 2025;12:100145. PMID: 40979181.

[33] Pfaffenseller B, Schneider J, De Azevedo Cardoso T, Simjanoski M, Alda M, Kapczinski F, et al. Self-assessment and rest-activity rhythm monitoring for effective bipolar disorder management: A longitudinal actigraphy study. International Journal of Bipolar Disorders. 2025;13(1):34. PMID: 41313566.

[34] Box GEP, Cox DR. An Analysis of Transformations. Journal of the Royal Statistical Society Series B (Methodological). 1964;26:211–52.

[35] Yeo IK, Johnson RA. A New Family of Power Transformations to Improve Normality or Symmetry. Biometrika. 2000;87(4):954–9. Available from: http://www.jstor.org/stable/2673623.

[36] Scott J, Colom F, Young A, Bellivier F, Etain B. An evidence map of actigraphy studies exploring longitudinal associations between rest-activity rhythms and course and outcome of bipolar disorders. International Journal of Bipolar Disorders. 2020 12;8. PMID: 33258017.

[37] Panchal P, de Queiroz Campos G, Goldman DA, Auerbach RP, Merikangas KR, Swartz HA, et al. Toward a Digital Future in Bipolar Disorder Assessment: A Systematic Review of Disruptions in the Rest-Activity Cycle as Measured by Actigraphy. Frontiers in Psychiatry. 2022 5;13. PMID: 35677875.

